# IMPROVE-IT2: Implementing non-invasive circulating tumor DNA analysis to optimize the operative and postoperative treatment for patients with colorectal cancer – Intervention Trial 2: Statistical analysis plan

**DOI:** 10.1101/2024.06.25.24309390

**Authors:** Kåre Andersson Gotschalck, Uffe Heide-Jørgensen, Lene Hjerrild Iversen, Claus Lindbjerg Andersen

## Abstract

**Background:** Surveillance after curative intended surgery and adjuvant chemotherapy for localized colorectal cancer (CRC) aim at detecting recurrence sufficiently early to allow efficient treatment. Minimally invasive blood-based analysis of circulating tumor DNA (ctDNA) has the potential to identify patients with microscopic residual disease early after surgery and adjuvant chemotherapy for CRC, with a median lead time of 9 months compared to standard-of-care CT-based surveillance. The IMPROVE-IT2 study is a randomized controlled trial investigating the effect of ctDNA-guided surveillance compared to standard-of-care CT-based surveillance in high-risk stage II and stage III CRC patients after curative intended surgery and adjuvant chemotherapy. The primary trial endpoint is the fraction of recurrence patients receiving curative intended surgery or local metastasis directed treatment within 3 years after primary surgery for CRC. Secondary endpoints include time-to-clinical-recurrence, overall survival, quality-of-life, and quality-adjusted life-years cost-effectiveness.

**Objective:** To outline a pre-determined statistical analysis plan (SAP) before patients have completed the first year of follow-up after primary surgery.

**Methods:** The SAP describes the IMPROVE-IT2 study design and endpoints, the randomization procedure, sample size estimation, and the specific statistical procedures and methods for analyzing efficacy outcomes. Health economics and quality of life analyses are not included in this SAP but will be analyzed separately. The SAP outlines the planned primary analyses, subgroup analyses, and a range of specific sensitivity analyses. To avoid bias, the final analyses of the IMPROVE-IT2 trial will adhere to the SAP. The SAP was approved after end of recruitment and before completion of the first 12 months of follow-up after primary surgery.

**Trial Registration:** Clinicaltrials.gov NCT04084249, registered in April 2019.

## 1. INTRODUCTION

### 1.1 PREFACE

Death of colorectal cancer (CRC) is primarily associated to distant metastases, present at the time of diagnosis or appearing later following a cancer-free period. At time of diagnosis, 75% of patients present with non-metastatic disease, for which standard of care is curative intended surgery[1]. Following surgery 15-20% of patients experience recurrence of which 70-90% are detected within 3 years of surgery[2–9]. Due to the high recurrence risk, adjuvant chemotherapy is standard of care for high risk UICC stage II and UICC stage III patients[10], but still approximately 25% experience recurrence[11]. Extending the duration of adjuvant chemotherapy offers no further reduction in recurrence rates[12–15]. Consequently, the only alternative option after adjuvant chemotherapy is surveillance aimed at detecting recurrence sufficiently early to allow efficient treatment[16–18].

The current surveillance strategy in Denmark consists of computed tomography (CT) scans of thorax and abdomen at postoperative months 12 and 36, and colonoscopy every fifth year until age 75 years. Increasing intensity of fixed interval CT based surveillance for stage II-III CRC has not been found to improve survival[5, 19, 20]. However, a larger proportion of patients receive curative intended surgery for recurrence with intense surveillance[5].

Methods to identify patients with microscopic residual disease after adjuvant chemotherapy are highly needed[21]. Minimally-invasive blood-based analysis of circulating tumor DNA (ctDNA) has this potential, since patients with positive ctDNA following adjuvant chemotherapy possess a risk of recurrence of close to 100%, and ctDNA negative patients a risk as low as 10%[22–24]. Also, longitudinal ctDNA analysis detect recurrence with an average lead time of ∼9 months compared to standard-of-care CT imaging[22, 23].

The IMPROVE-IT2 randomized trial will explore the clinical utility of ctDNA guided surveillance following primary surgery and postoperative adjuvant chemotherapy for high-risk stage II and stage III CRC[25]. Specifically, ctDNA guided surveillance compared to standard-of-care CT-based surveillance. In concordance with clinical trial requirements, the study was prospectively registered (clincaltrials.gov, NCT04084249). The trial completed the target accrual in July 2023 with randomization of the final participant in February 2024. All participants will be followed up for at least 36 months after primary surgery.

### 1.2 PURPOSE OF THE ANALYSES

These analyses will assess the clinical utility of ctDNA-guided recurrence surveillance strategy in comparison with the standard-of-care CT-based approach. The statistical analysis plan will elaborate on the analyses targeting the endpoints mentioned below.

## 2 STUDY OBJECTIVES AND ENDPOINTS

### 2.1 STUDY OBJECTIVES

#### 2.1.1 Primary Objective

The primary objective is to investigate if ctDNA guided post-operative surveillance to guide radiological assessments will result in a higher fraction of recurrence patients receiving curative intended or local metastasis-directed treatment for CRC recurrence as compared to current Danish surveillance strategy within 3 years after primary surgery.

#### 2.1.2 Key Secondary Objective

##### Oncological

- Key Secondary objective 1 (S1) To investigate if ctDNA guided surveillance is associated with a shorter time from randomization to clinical recurrence (TTCR) than standard-of-care surveillance.
- Key Secondary Objective (S2) To investigate if 3- and 5-year overall survival (OS) after surgery is non-inferior with ctDNA guided surveillance compared to current standard-of-care CT based surveillance. To take immortal time, typically 4-8 months, from the time of surgery until randomization into account, overall survival will be estimated with two different starting points of follow-up : 1) from time of surgery and 2) from time of randomization. For both starting points, OS will be assessed at 3- and 5-years after surgery.

##### Quality of Life

- Key Secondary objective 3 (S3): To compare the quality of life (EORTC QLQ-C30 and EQ-5D-L5), fear of cancer recurrence inventory (FCRI), and impact of events scale for cancer (IES-C) for patients following ctDNA guided and the standard-of-care surveillance.

#### 2.1.3 Secondary Objectives

- Secondary objective 4 (S4) To assess the cost-effectiveness of ctDNA-guided and standard-of-care surveillance, to provide decision support for clinicians and other decision makers.
- Secondary objective 5 (S5) To assess, and compare, the protocol adherence rates (AR) for patients following ctDNA guided and standard-of-care surveillance.
- Secondary objective 6 (S6) To assess if the time to molecular recurrence (TTMR), i.e., detection of ctDNA- and/or elevated level of Carcinoembryonic antigen (CEA) are similar, for patients following ctDNA guided and patients following standard-of-care surveillance.
- Secondary objective 7 (S7) To describe any changes in quality of life (EORTC QLQ-C30 and EQ-5D-L5), fear of cancer recurrence inventory (FCRI) and impact of events scale for cancer (IES-C) from time of positive ctDNA until time of clinical verified recurrence.
- Secondary objective 8 (S8) To investigate if ctDNA growth rate analyses, performed on blood samples collected within a short interval (2-3 weeks apart), can stratify ctDNA positive patients into groups of fast and slow growing tumors.
- Secondary objective 9 (S9): To compare the rate of clinical recurrence detection during the time-intervals: randomization-12 months after surgery, 13-24 months after surgery, and 25-36 months after surgery for patients with ctDNA guided and standard-of-care surveillance.
- Secondary objective 10 (S10). To compare the cumulative incidence of clinical recurrence at 12 months, 24 months and 36 months after surgery for patients with ctDNA guided and standard-of-care surveillance.

### 2.2 DEFINITIONS OF PREDICTOR VARIABLES, CO-VARIATES AND ENDPOINTS

#### 2.2.1. Primary endpoint

- Fraction of recurrence patients receiving intended curative resection or local treatment aiming at complete tumor destruction, as defined prospectively by the Endpoint committee, within 3 years after surgery.

#### 2.2.2 Secondary endpoints

- Time to clinical recurrence (TTCR)
- Overall survival at 3 and 5 years after surgery (3-year OS and 5-year OS)
- Time to molecular recurrence (TTMR) by either ctDNA or CEA
- Clinical recurrence rate at time-intervals: randomization-12 months, 13-24 months, and 25-36 months.
- Cumulative incidence function of clinical recurrence at 1, 2, and 3 years (1, 2, and 3-year CIF)
- Quality of Life (QoL) by use of European Organization for Research and Treatment of Cancer Quality of Life questionnaire, Core 30 (EORTC QLQ-C30), version 3.0, using global health status and functional scales only (17 items)
- Fear of Cancer Recurrence Inventory (FCRI) (42 items)
- Impact of Events Scale Cancer (IES-C) (15 items)
- European Quality of life – 5 Dimensions (EQ-5D-5L) (5 items)
- Adherence rate (AR)
- Cost-effectiveness (CE)

#### 2.2.3 Safety Endpoints

- Frequency and severity of adverse events (AE) in relation to the per protocol blood draws.
- Frequency of allergic reactions to contrast material in relation to PET/CT imaging.

#### 2.2.4 Definition of end-points

- Definition of curative or palliative intended treatment of recurrence: An Endpoint committee appointed by the trial office will prospectively and independently assess the local site multidisciplinary team (MDT) conference notes and medical records of all recurrence patients (both arms) and decide if the planned treatment is curative intended or palliative. The decision will not be provided to the local sites or otherwise influence the actual treatment delivered.
- Overall survival from time of surgery is the outcome of primary interest and is calculated as the time from surgery to death from any cause. Due to immortal time from time of surgery until time of randomization (up till 8 months) an additional analysis will be performed calculating overall survival from time of randomization to death from any cause.
- Time to clinical recurrence is calculated from randomization until detection of loco-regional recurrence or distant metastases, or death from colorectal cancer.
- Time to molecular recurrence is calculated from randomization until detectable ctDNA with censoring at time of death or end of follow-up.
- Clinical recurrence rate is defined as total number of clinical recurrences detected divided by the total follow-up time during the specific time-interval.
- Cumulative incidence function of clinical recurrence is calculated treating clinical recurrence as event and death as competing event with censoring at end of follow-up.
- Quality of Life is assessed by the questionnaires: QLQ-C30 (Appendix D), fear of cancer recurrence inventory, FCRI (Appendix E), impact of events scale cancer, IES-C (Appendix F), European Quality of life – 5 Dimensions, EQ-5D-5L (Appendix G).
- Adherence rate assessed by the proportion of patients adhering 100% to the protocol. Protocol adherence is for both arms defined as attendance to all planned diagnostic surveillance according to the protocol and no extra planned surveillance (diagnostic examinations for clinical indications allowed).
- The cost-effectiveness analysis will be carried out from a health care perspective and the health outcome measure in the cost-effectiveness analysis will be the total quality adjusted life years per group.

## 3 STUDY METHODS

### 3.1 STUDY DESIGN

IMPROVE-IT2 is a randomized multicenter trial comparing the effect of ctDNA guided post-operative surveillance and standard-of-care CT scan surveillance[25]. The trial design is a parallel group study with 1:1 allocation. The trial profile is illustrated as a flow chart in figure 1. The full protocol (see supplementary file) is consistent with current Standard Protocol Items: Recommendations for Interventional Trials (SPIRIT) guidelines[26, 27].

**Figure 1.**
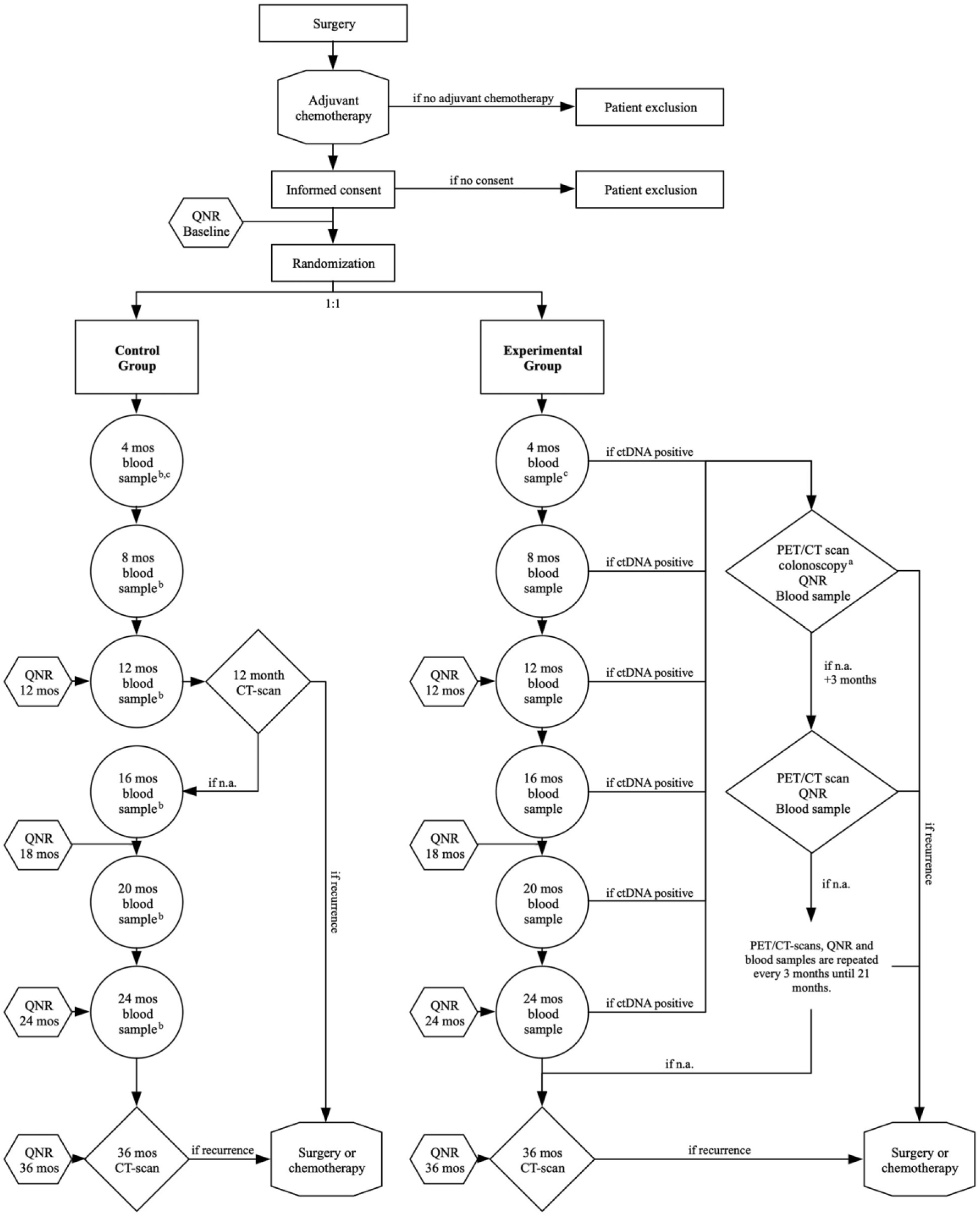
Trial profile. ctDNA: circulating tumor DNA; PET/CT: Flour-Deoxy-Glucose Positron Emissions Tomography scan; FCRI: fear of cancer recurrence inventory; fear of cancer recurrence inventory; IES-C: Impact of Event Scale - Cancer; Mos: months; N.a.: nothing abnormal; QNR: questionnaires. A colonoscopy is performed in case of ctDNA blood sample and a PET-CT scan with no evidence or suspicion of residual disease or recurrence. Blood samples from the control group are not analyzed until end of study, but serve to enable post-trial comparison of oncological outcomes for the two groups stratified for ctDNA-status. A 4-month post-operative blood sample is taken in case of 3 months adjuvant chemotherapy regime. In case of 6 months adjuvant chemotherapy the 8-month post-operative blood sample will serve as the first blood sample. QNR: At baseline (randomization) and post-operative months 12-, 18-, 24- and 36-months patients complete the QoL questionnaires including EORTC QLQ-C30, EQ-5D-5L, FCRI, and IES-C. Further, ctDNA positive patients will complete the QNR questionnaires before each PET-CT scan.[25]

### 3.2 STUDY POPULATION AND ELIGIBILITY CRITERIA

In total 11 of 17 surgical centers in Denmark are participating. The trial includes 359 patients with exclusion of 60 patients prior to randomization, leaving 299 patients with resected CRC and adjuvant chemotherapy randomized to either ctDNA guided or standard-of-care CT-based surveillance. Eligibility criteria are the following:

#### Inclusion criteria

- Colon or rectal cancer, tumor stage III (pT1-4N1-2,cM0) or stage II high risk (pT4N0,cM0 and pT3N0, cM0 with either of the risk factors (<12 examined lymph nodes, anastomotic leakage, emergency surgery, signet ring adenocarcinoma) described in the national guideline for adjuvant therapy to stage II cancer)
- Received intended curative resection and found eligible for adjuvant chemotherapy

#### Exclusion criteria

- Adjuvant chemotherapy not initiated
- Synchronous colorectal and non-colorectal cancer diagnosed perioperatively (except skin cancer other than melanoma)
- Other cancers (excluding CRC or skin cancer other than melanoma) within 3 years from screening for eligibility

### 3.3 RECUITMENT

Patients who meet the eligibility criteria are approached at the treating surgical department. The patients are given written project information and oral information by a trained health care professional. As the project involves genomic sequencing, the participants are offered genetic counseling before obtaining written informed consent. Informed consent is given on a voluntary basis and must be obtained prior to commence of any study-related procedure. The consent may be withdrawn at any time without having any impact on current or future treatment.

### 3.4 RANDOMIZATION

Randomization is performed using a concealed centralized web-based randomization service with data linked to the Research Electronic Data Capture (REDCap) database [28, 29]. A block randomization is set up to stratify by tumor location, pT- and pN -category, and standard-of-care surveillance program intensity, as these factors are associated with risk, location and prognosis of recurrence, and timing of recurrence detection.

### 3.5 INTERVENTIONS

#### The experimental group

ctDNA analysis will be performed every 4 months postoperatively (4, 8, 12, 16, 20 and 24). At time of first positive ctDNA, patients undergo a whole-body FDG-PET/CT scan for radiological assessment and a colonoscopy. If the initial assessment is without evidence of recurrence or another cancer, patients will be offered a colonoscopy and high-intensive radiological surveillance with FDG-PET/CT scans every 3 months, until recurrence detection or 21 months have passed.

#### The control group

Patients will undergo surveillance according to current Danish Guidelines with CT scans at months 12 and 36 postoperatively and colonoscopy every 5 year until age 75. Longitudinal blood samples will be collected at same time-points as in the experimental group but not analyzed until end of trial.

Both patient groups complete QoL questionnaires at baseline (prior to being informed about randomization allocation and start of surveillance), and at months 4, 12, 18, 24 and 36 months. The questionnaire includes EORTC QLQ-C30 ver. 3.0 [29], EQ-5D-5L[30,31], FCRI[32], and IES-C[33]. Before every FDG-PET/CT scan in ctDNA positive patients, the patients also complete the FCRI questionnaires.

## 4. SAMPLE SIZE

Sample size assessment is made for the primary outcome and, additionally, for the key secondary outcomes.

### 4.1 Assessment of sample size needed to meet the primary objective

The recurrence rate in this population is 25%, and we assume this to be equal within 36 months after primary surgery in both arms. Primary endpoint of the study is the fraction of recurrence patients receiving curative-intent resection or local treatment. With standard-of-care surveillance, approximately 15% of recurrence events are eligible to curative-intent treatment. We assume this fraction can be increased by a factor of 3 (to 45%) in the experimental arm. To achieve a power of 80% to detect the difference, at a 5% significance level, a sample size of 33 recurrences (132 patients with a 25% recurrence rate) in each arm is needed. Assuming a drop-out rate of 14%, a total of 306 patients needs to be included.

The sample size estimation above is based on the assumption that the recurrence rate will be equal within 36 months after primary surgery in both arms. However, as the intervention is a different surveillance strategy, there could theoretically be different proportions of recurrences at 36 months in the intervention and control surveillance arm. Therefore, a sensitivity analysis will be performed which will compare the fraction of patients with curative-intent treatment for recurrence relative to the entire population (recurrence and non-recurrence patients) for patients in the experimental and the standard-of-care surveillance arms.

KEY SECONDARY ENDPOINTS

### 4.2 Assessment of sample size needed to meet key secondary objective, time to clinical recurrence (TTCR) (S1)

Based on data of 141 patients included in a previous published *observational* study[30] with ctDNA samples taken serially after end of adjuvant chemotherapy and during standard of care CT-based recurrence surveillance, we can estimate the difference in the observed time to recurrence and the theoretical time to recurrence with ctDNA guided surveillance. This under the assumption that patients would have recurrence detected at time of first positive ctDNA sample in an interventional setting. Using a generalized gamma accelerated failure time regression (best fitted model), we estimate a time ratio of 0.25 (95% CI: 0.14-0.42) for recurrence at time of ctDNA positive sample compared to the actual time of clinical recurrence in the cohort. A time ratio <1 results in a proportional shift to the left in the Kaplan-Meier curve (earlier recurrence detection), see figure 2. We therefore believe our sample size is sufficient to demonstrate a shorter time to recurrence with ctDNA-guided surveillance.

**Figure 2.**
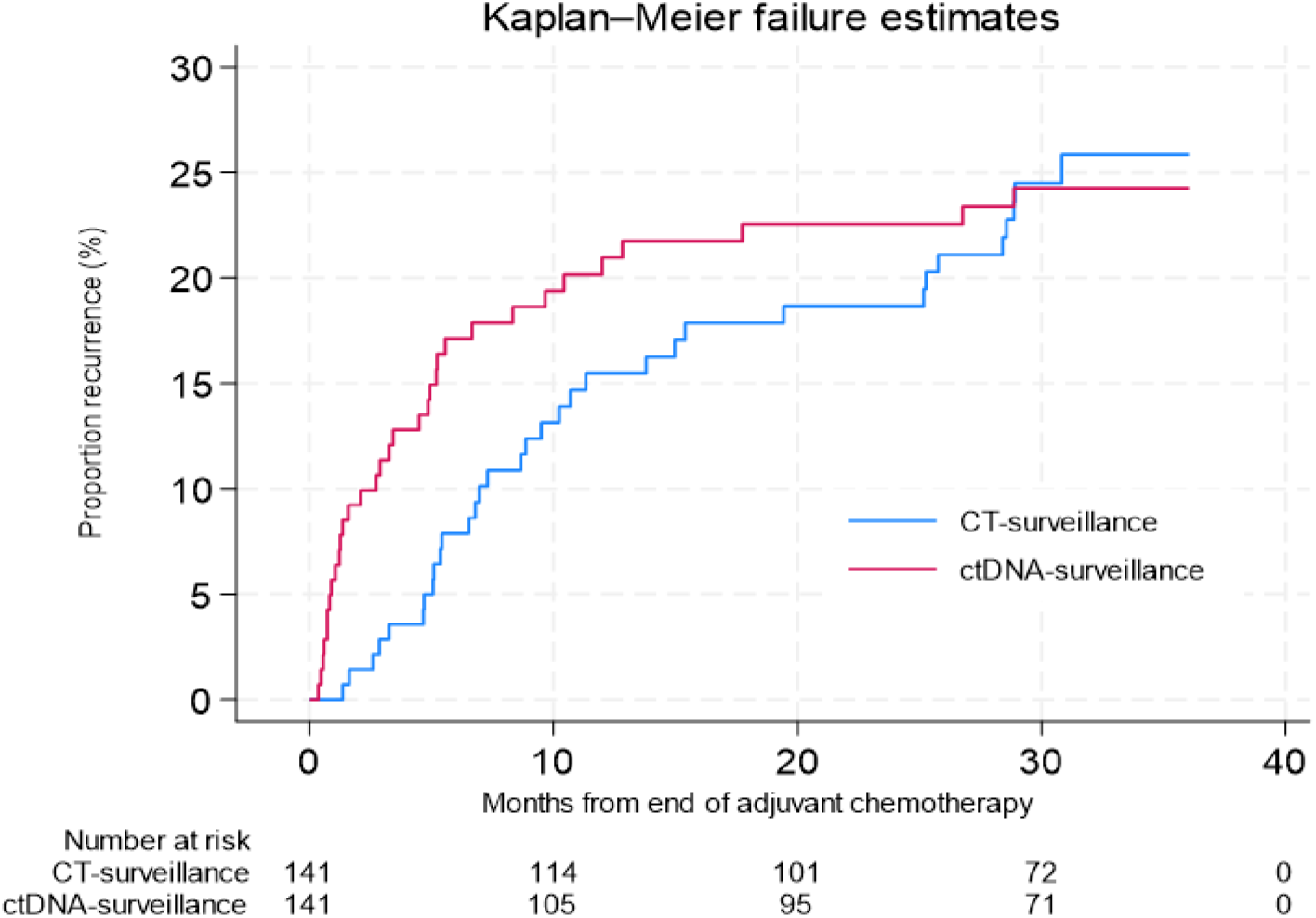
Kaplan-Meier failure estimates of recurrence detection at the actual time of clinical recurrence (“CT-based surveillance”) compared to the time-point of first ctDNA positive sample after end of adjuvant chemotherapy (“ctDNA-surveillance”).

### 4.3 Assessment of sample size needed to meet key secondary objective, overall survival (S2)

In accordance with estimates based on data from a recent published Danish nation-wide cohort study [11], the 3-year overall survival following surgery for stage II-III CRC among patients who receive adjuvant chemotherapy is estimated at 92%. A non-inferior limit of 8%-point have been used previously in ctDNA trials [31]. If there is truly no difference between the 3-year OS in standard-of-care CT based surveillance arm and the experimental ctDNA-guided surveillance arm (92% in both groups), then 286 patients (143 in each arm) are required to be 80% sure that the upper limit of a one-sided 95% confidence interval (or equivalently a 90% two-sided confidence interval) will exclude a difference in favor of the standard group of more than 8%-point.

### 4.4 Assessment of sample size needed to meet key secondary objective, Quality of Life (S3)

A mean global health-status QoL difference of 10 points is defined to be the threshold of clinical relevance[32, 33]. Assuming a mean global health-status/QoL score of 73 with a standard deviation of 23 [33] then with no difference between experimental and control arm, 91 patients in each arm are required to show with a power of 90% that the lower limit of a one-sided 95% confidence interval will be above the non-inferiority limit of −10.

## 5 GENERAL CONSIDERATIONS

### 5.1 TIMING OF ANALYSES

The final analyses of the primary objective and key secondary objectives will be performed when all study subjects have completed the 36 months post-operative CT scan, have experienced recurrences or have dropped out. Patients in the experimental arm enrolled in PET/CT scan every 3 months after ctDNA positive sample will be followed for minimum 36 months postoperative or until the final PET/CT (up to 24 months after ctDNA positivity), but the recurrence status at 36 months post-operative will be used for analysis. CT scans performed from 34-38 months postoperative will be classified as the final 36 months surveillance CT scan in both arms. In case of multiple CT scans during this timeframe, the latest will be included. The reason for allowing this interval is based on the clinical reality that CT scans planned on a fixed time-point (i.e., 36 months post-surgery) are often scheduled and performed within a short timeframe around this exact time-point.

The overall survival analyses will be performed when 5 years have passed for all study subjects from time of surgery. Overall survival from time of surgery is the outcome of primary interest. However, due to immortal time from time of surgery until randomization, start of follow-up for the overall survival analyses will be considered at two time-points: 1) at the time of surgery for both arms and, additionally, 2) at the time of randomization for both arms.

### 5.2. ANALYSES POPULATION

#### 5.2.1 Full analyses population

Will include all study subjects who were randomized (Excluding patients withdrawing consent)

#### 5.2.2 Per protocol “clinical objectives” analyses

Protocol deviations that would exclude a patient from the per protocol analysis are described in detail in section 4.2. In summary, the analyses will include all study subjects except:

##### Experimental arm

patients that had one or more non-protocolled *planned* CT surveillance scan without clinical indication (the planned 36-month CT-scan and extra scans for clinical reasons allowed).

##### Control arm

Patients that prior to randomization had more intensive radiological surveillance *planned* than the standard-of-care surveillance protocol (extra scans for clinical reasons allowed).

### 5.3 COVARITES AND SUBGROUP

Important covariates that will be adjusted for in the primary analysis are:

- Age (continuous variable)
- Sex

Block-randomization is made on the following variables: T category (T4 vs. T1-3), N category (N0-1 vs N2), primary tumor location (rectum/colon), and high vs. low (CT 12 and 36 months, only) intensity surveillance centers. Therefore, no adjustment will be made for these co-variates in the primary analysis.

Exploratory subgroup-specific ***summary*** statistics will be presented as forest plot figures.

### 5.4 MISSING DATA

The frequency of missing baseline variables and outcome (either binary or time to event) is expected to be minimal, therefore no formal imputation method will be performed. However, any patient lost to follow-up within the first three years after surgery without recurrence, will be coded as no recurrence for the primary efficacy outcome and censored for the time to event outcomes.

### 5.5 INTERIM ANALYSIS, DATA MONTORING AND ENDPOINT COMMITTEE

#### 5.5.1 Purpose of interim analyses

Interim analyses are conducted to demonstrate the feasibility and safety of recurrence detection by ctDNA guided surveillance; thus, the aim is to access safety of ctDNA-guide surveillance in recurrence detection. Interim analyses are conducted every month and until all patients has concluded the first 12 months of post-operative follow-up (or detected recurrence). The results of interim analyses at the time of the statistical analysis plan (>90% concluded 12 months follow-up) showed that ctDNA guided surveillance did not deviate from standard recurrence detection.

#### 5.5.2 Data monitoring

A Data and Safety Monitoring Board (DSMB) was established to monitor study conduct and progress. The DSMB review on a weekly basis unblinded data to examine the quality of ctDNA analyses, ctDNA results and compliance to the allocated surveillance program. In addition, an IMPROVE-IT2 Steering committee with an external advisory board was established to monitor overall study progress and progress on trial deliverables with meeting every 6 months during the trial period.

#### 5.5.3 Endpoint committee

An Endpoint committee was established to independently and uniformly classify recurrence status and intent of recurrence treatment. The committee is consulted on all cases of recurrences to established 1) if a recurrence is detected and 2) if the *planned* treatment should be classified as curative intended recurrence treatment (primary endpoint) or palliative treatment. To avoid retrospective classification of the study endpoint, the evaluations of the Endpoint committee is made prospectively during the study, allowing for immediate clarification from the treating centers in case the intent of the planned recurrence treatment is unclear or questionable.

### 5.6 MULTI-CENTRE STUDIES

The data from participating centers in this protocol will be combined, so that an adequate number of patients will be available for analysis. Centers will in subgroup analysis be stratified by centers with high-vs. low-intensity standard-of-care surveillance programs to account for potential effect of different standard-of-care surveillance programs.

### 5.7 MULTIPLE TESTING

No formal multiplicity testing adjustment will be performed. However, the outcomes are clearly ranked by degree of importance (primary, and secondary), and a limited number of pre-specified subgroup analyses will be conducted.

## 6 SUMMARY OF STUDY DATA

All continuous variables will be summarized using the following descriptive statistics: number (non-missing sample size), mean, standard deviation, median, maximum and minimum. The frequency and percentages (where the denominator consist of sample availability for each associated variable) of observed levels will be reported for all categorical measures. All summary tables will be structured with a column for each surveillance arm in the order (Control, Experimental) and will be annotated with the total population size relevant to that table/surveillance strategy, including any missing observations.

### 6.1 SUBJECT DISPOSITION

CONSORT diagram: a trial profile will illustrate patients’ progression through the study from initial screening for eligibility to completion of follow-up (see Figure 3)

**Figure 3.**
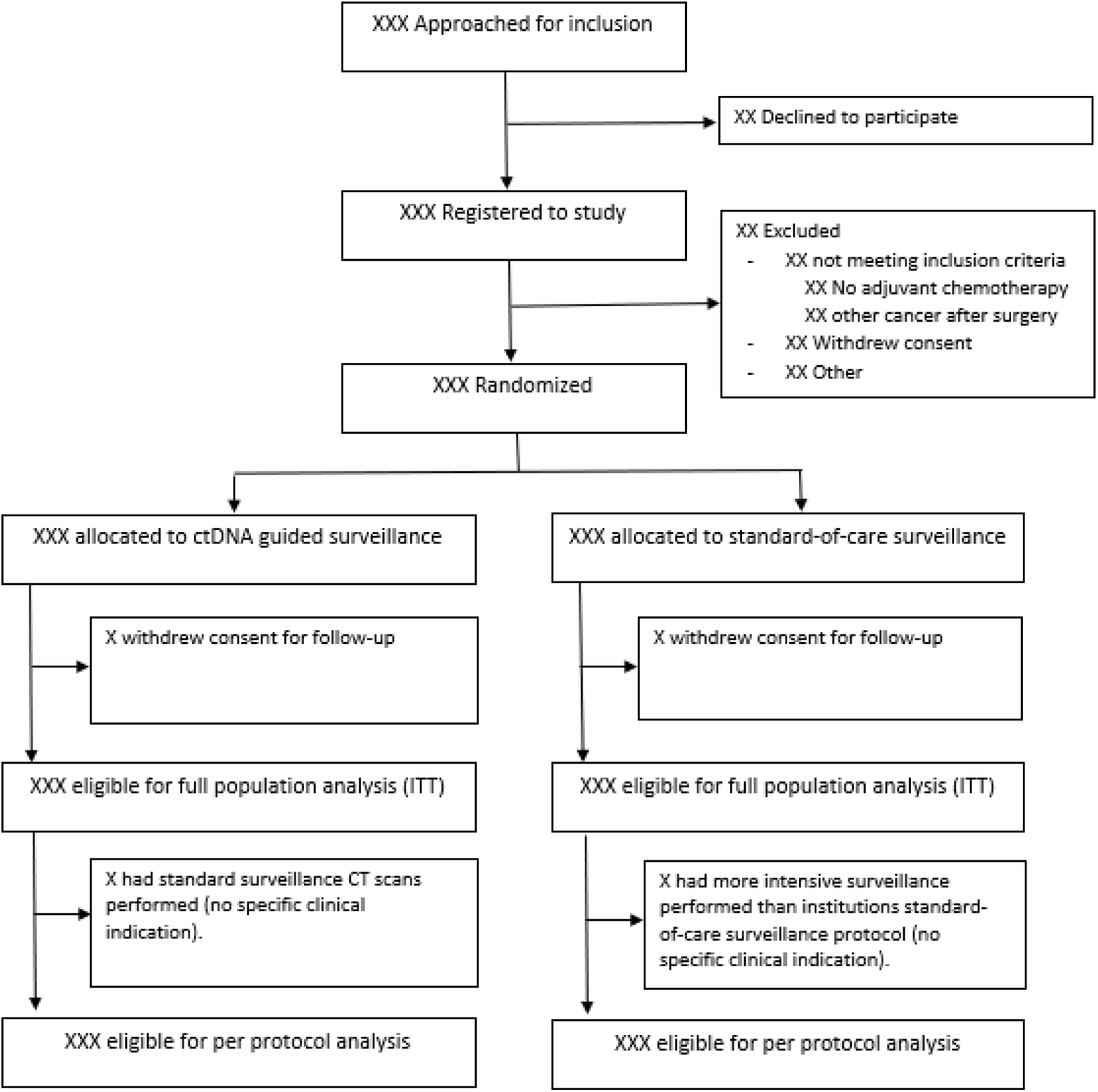
CONSORT diagram

### 6.2 PROTOCOL DEVIATIONS

Major deviations that would exclude a subject from the per protocol analysis in the experimental arm include shift from ctDNA-guided follow-up to standard-of-care CT based surveillance, with at least one standard CT surveillance scan performed before the 36 months end-of-study CT scan. In the standard-of care surveillance arm with, a planned imaging schedule more intensive than the institution’s standard-of-care surveillance program would exclude the study subject from the per protocol analysis. Additional imaging for clinical reasons (E.g., due to symptoms) is allowed in both arms and not considered a protocol violation.

### 6.3 DEMOGRAPHIC AND BASELINE VARIABLES

Patient baseline characteristics, type of primary surgery, pathology details of the resected primary tumor, adjuvant chemotherapy (administered drugs, duration of chemotherapy, start and stop dates, number of cycles, reasons for stopping chemotherapy) will be summarized by study arm using mean (sd) and/or median (range) for continuous variables and frequency (proportion) for categorical variables. Since any differences between randomized groups at baseline could only have occurred by chance, no formal significance testing will be carried out.

Baseline as well as 12 months QoL data will be presented in a separate publication prior to publication of the main trial data.

## 7 EFFICACY ANALYSES

### 7.1 GENERAL CONSIDERATIONS

The analysis principles are as follows:

1. Efficacy analyses will be conducted both on an intention-to-treat (ITT) and per protocol basis (PP) but the ITT analysis remains the gold standard
2. All randomized patients will be analyzed in the group to which they were assigned regardless of protocol violations. The only exception will be patients whose consent to use their data in the analysis is withheld or withdrawn.
3. All tests will be two-sided with a nominal significance level of 5%.
4. Effect of the surveillance strategy will be estimated as difference in proportions, means, odds ratio, hazard ratio or time ratio along with their 95% confidence interval (CI) and will be reported for all outcomes.
5. Subgroup analyses will be carried out irrespective of whether there is a significant effect of surveillance strategy on the primary outcome.
6. P values will not be adjusted for multiplicity. However, a limited number of subgroup analyses are pre-specified.
7. All summary tables will be annotated with the total population size relevant to each surveillance group.
8. P values ≥ 0.001 will be reported to three decimal places; P values less than 0.001 will be reported as ‘< 0.001’. The mean, SD and any other statistics other than quantiles will be reported to one decimal place greater than the original data. Quantiles, such as median, or minimum and maximum will use the same number of decimal places as the original data. Estimated parameters, not on the same scale as raw observations (e.g., regression coefficients), will be reported to two significant figures.
10. Analyses will be conducted primarily using SAS, version 9.4 or later, R 3.4.1 or later or STATA version
11. or later.

### 7.2 PRIMARY EFFICACY ANALYSIS

The primary outcome analysis of the fraction of patients with curative intended treatment for recurrence will be based on the intention-to-treat population. A sensitivity analysis will be performed using the per-protocol population.

The distribution of recurrence patients with and without curative-intended treatment stratified on surveillance arm will be presented in a 2×2 table. A log-binominal regression analysis will be used to estimate the relative risk of curative-intended recurrence treatment in the experimental arm compared to the standard-of-care arm.

A sensitivity analysis will be performed which will compare the fraction of patients with curative-intent treatment for recurrence relative to the entire population (recurrence and non-recurrence patients) for patients in the experimental and the standard-of-care surveillance arms (see section 4.1).

### 7.3 SECONDARY EFFICACY ANALYSIS

#### Time-to clinical recurrence

Curves displaying the cumulative incidence proportion of recurrences in either arm will be constructed using the Aalen-Johansen estimator for visualization. We expect the cumulative incidence of recurrence at 36 months post-operative to be similar in each arm but expect the time-points of recurrence detection during the study period to be different (earlier recurrence detection in the ctDNA-guided arm). We do not expect proportional hazards, and a robust measure which does not rely on a specific assumption will be generated to compare difference in time to clinical recurrence between the two surveillance arms.

#### Time to molecular recurrence

Curves displaying the cumulative incidence proportion of *molecular* recurrences (ctDNA positive) in either arm will be constructed using the Aalen-Johansen estimator for visualization. We expect the cumulative incidence of molecular recurrence at 36 months post-operative to be similar in each arm with similar time-points of molecular recurrence detection during the study period. We expect the proportional hazard assumption to be valid but will use similar statistics as time to clinical recurrence analyses. In the control arm, the median (range) lead-time from molecular to clinical recurrence detection will be presented.

#### Overall survival

will be described using the product limit method of Kaplan-Meier. Survival curve distribution differences between arms will be tested using log-rank test. Hazard ratios and corresponding 95% CI will be computed using the Cox proportional hazard model. The validity of the proportional hazards assumption will be tested using Shoenfeld residuals plots and corresponding test statistics. To account for take immortal time from the time of surgery until randomization, we will analyze overall survival both from time of surgery as well as from time of randomization.

### 7.4 EXPLORATORY EFFICACY ANALYSES

1. Subgroup: The primary endpoint and the key secondary endpoints will be assessed on a limited number of subgroups. The objective of this analysis is to assess potential differences in surveillance strategies effects between subsets of patients, through an interaction term effect. Evidence of heterogeneity of surveillance strategies effects among subgroups will be demonstrated by the level of statistical significance of the interaction term between treatment group and subgroup using either a log-binomial regression (for a binary outcome), Cox regression (overall survival) or accelerated time failure model (time to recurrence) using a threshold of significance of 0.05. The subgroup analyses will remain exploratory and hypothesis generating given the study is not specifically powered to test any effect modification. The following factors will be investigated:

a. Participating center (high vs low-intensity standard-of-care surveillance)
b. T category (T1-T3 vs T4)
c. Sex (male vs female)
d. Age (dichotomized (<70 vs ≥70 years))
e. Primary tumor site (colon vs rectum)
f. Lymph Nodes Examined (> 12 vs ≤12)
g. N category (N1 vs N2)
2. Exploratory analyses of the association between preoperative ctDNA levels both as a categorical variable (positive or negative) and as continuous variable (mutant allele fraction) with the risk of recurrence will be performed. Furthermore, we will examine the theoretical effect changing the level for calling the post-operative ctDNA positive could have on the time to recurrence and the false-positive/false-negative rates.

### 7.5 SAFTY ANLYSES

Given no treatments that are beyond standard of care were administered in this study, no safety analysis will be performed.

## 8 CONCLUSION

We have developed a statistical analysis plan (SAP) for the IMPROVE-IT2 study[25]. This plan will be followed to ensure high-quality standards of internal validity to minimize analysis bias. The SAP was approved and signed off by the study chairs and biostatistician on June 14, 2024

## 9 LIST OF TABLES AND FIGURES

A list of proposed tables and figures are displayed in this section.

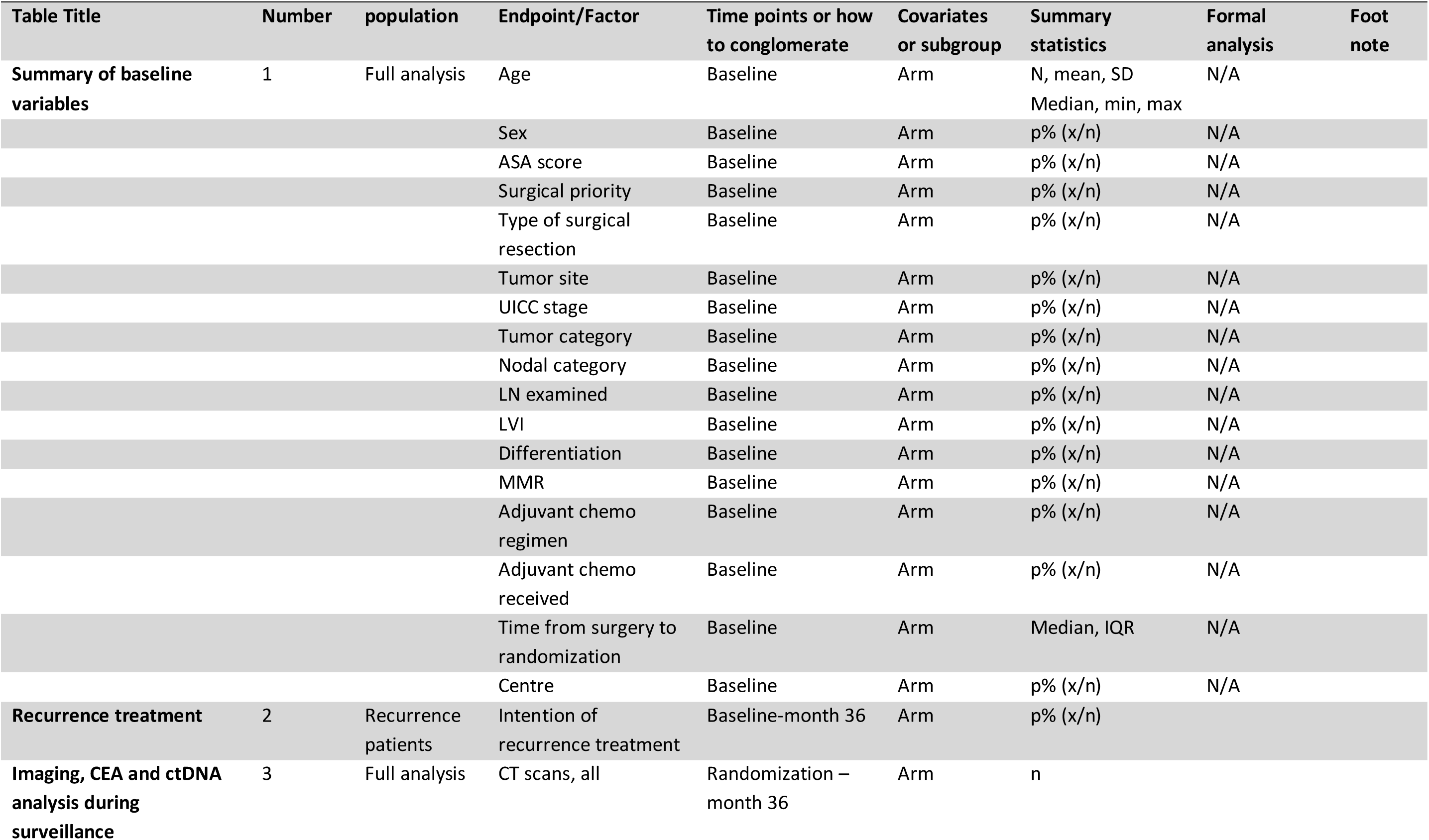
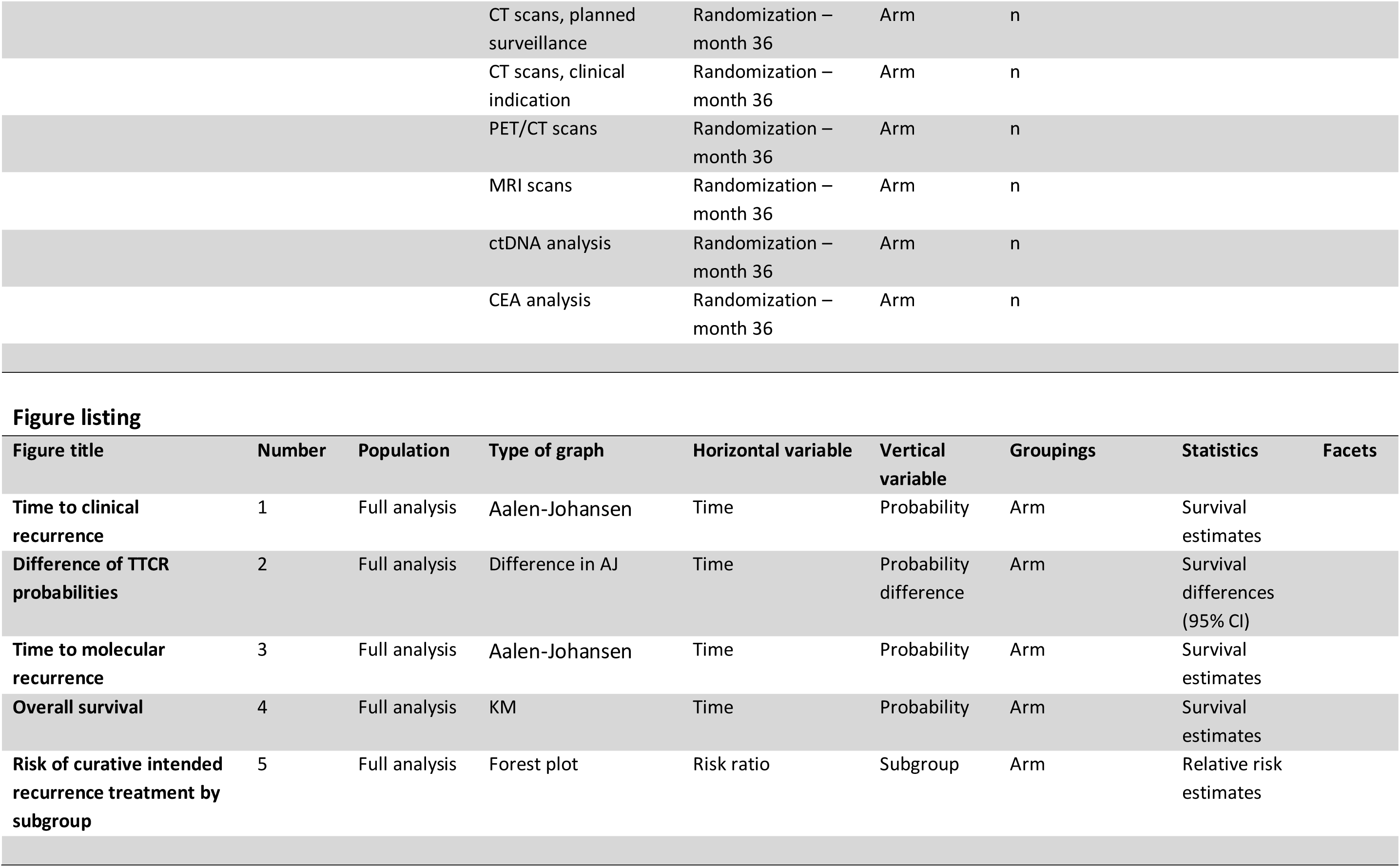
Table listing.

## Data Availability

Danish legislation and the conditions of the ethical approval for the study preclude public sharing of primary data and materials.

